# Early prediction of childbirth-related posttraumatic stress disorder symptoms

**DOI:** 10.64898/2026.03.18.26348512

**Authors:** Hadas Allouche-Kam, Isha Hemant Arora, Mary Lee, Francine Hughes, Sharon Dekel

## Abstract

Childbirth-related posttraumatic stress disorder (PTSD) is an underrecognized maternal morbidity. We tested whether obstetric complications increase risk for later PTSD symptoms through acute peritraumatic distress and early postpartum trauma symptoms. In a prospective cohort of 667 women, acute distress was assessed at 1.44 days postpartum, PTSD symptoms at approximately 10 days and 2 months postpartum, and depression symptoms at 10 days; obstetric complications were abstracted from medical records. Structural equation modeling showed that obstetric complications were associated with greater acute distress (β=0.292, p<0.001), which predicted higher PTSD symptoms at 10 days postpartum (β=0.561, p<0.001); early symptoms predicted symptoms at 2 months (β=0.665, p<0.001). The direct path from obstetric complications to 2-month PTSD symptoms was not significant. A significant serial indirect effect was observed through acute distress and early PTSD symptoms (β=0.109, p<0.001), whereas depression was not a mediator. These findings support early screening for childbirth-related PTSD risk after complicated delivery.

## Introduction

Childbirth can be experienced as psychologically traumatic ^1^. According to DSM-5, exposure to a traumatic event involving perceived or actual sense of threat of death or serious physical injury can result in posttraumatic stress disorder (PTSD). Growing evidence indicates that a subset of women develops persistent, traumatic symptoms specifically linked to childbirth^1^, with up to 16.8% having clinical symptoms^1^, which are associated with heightened physiological reactivity to traumatic reminders, akin to patterns observed in veterans with PTSD^2^. Globally, rates of obstetric complications remain disproportionately high in low- and middle-income countries, where life-threatening delivery events continue to account for substantial maternal morbidity and mortality^3^, underscoring the importance of understanding both the medical and psychological sequelae of complicated childbirth.

According to psychiatric nosology, PTSD cannot be formally diagnosed until symptoms persist for at least one month. The dose-response model of PTSD posits a strong association between event magnitude and clinical outcome. Medically complicated childbirth involving maternal or infant acuity may provide early indicators of PTSD risk^4^; however, objective event characteristics alone may be insufficient to determine risk. Because peritraumatic (immediate) distress reactions, including emotional responses and subjective threat appraisals, are established risk factors for the development of PTSD^5,6^, we examined whether obstetric complications are associated with acute subjective distress during delivery and whether acute distress and early postpartum symptoms subsequently explain risk for later PTSD symptoms.

## Methods

A total of 667 women who recently gave birth at Massachusetts General Hospital were studied in a prospective longitudinal study through the third trimester and the Postpartum period. The study was approved by the hospital’s Human Research Committee. Maternal age was approximately 34, nearly half (40%) of participants were primiparous; and 43.03% experienced one or more obstetric complications (Table 1).

**Table 1.** Demographics and obstetrics information of the study sample.

| Variable | *n (%) / M (SD)* |
| --- | --- |
| **Demographic** |  |
| Maternal age at delivery | 33.86 (4.18) |
| Marital status |  |
| Married or living with partner | 631 (94.60%) |
| Single | 36 (5.40%) |
| Education |  |
| Formal college degree or higher | 583 (87.41%) |
| No formal degree | 84 (12.59%) |
| Parity |  |
| Primiparas | 259 (38.83%) |
| Multiparas | 408 (61.17%) |
| Gestation age (wks) | 39.20 (1.31) |
| **Mode of delivery** |  |
| Vaginal | 435 (65.22%) |
| Vaginal assisted | 40 (6.00%) |
| Scheduled Cesarean | 79 (11.84%) |
| Unscheduled or emergency Cesarean | 113 (16.94%) |
| **Obstetric Complications** |  |
| 3<sup>rd</sup> or 4<sup>th</sup> degree lacerations | 17 (2.55%) |
| Anesthetic complications | 2 (0.30%) |
| Coagulopathy | 2 (0.30%) |
| Failure to progress in 1<sup>st</sup>/2<sup>nd</sup> stage | 76 (11.39%) |
| Fetal intolerance of labor | 69 (10.34%) |
| Hemorrhage | 82 (12.29%) |
| Hypertensive complications of pregnancy | 23 (3.45%) |
| Hysterotomy extension to the vagina | 4 (0.60%) |
| Manual removal of retained placenta | 40 (6.00%) |
| Malpresentation | 33 (4.95%) |
| Placental abruption | 11 (1.65%) |
| Sholder dystocia | 6 (0.90%) |
| Suspected chorioamnionitis | 17 (2.55%) |
| Unplanned hysterectomy | 1 (0.15%) |
| NICU admission | 62 (9.30%) |

Note. Medical information derived from electronic medical records. Obstetric complications = indicators of maternal and fetal acuity related to labor and delivery. Hemorrhage defined as estimated blood loss $\geq 1000$ ml or indicated specifically in the medical records. NICU = Neonatal Intensive Care Unit admission. *n* = 667.

Acute distress experienced during or immediately after childbirth, reflecting responses such as fear, helplessness, and perceived subjective threat, was assessed in the immediate postpartum assessment, conducted primarily during maternity hospitalization stay, and occurring on average 35 hours postpartum (mean=1.44 days, standard deviation=1.38). Acute reactions were measured with the Peritraumatic Distress Inventory (PDI)^5^, an instrument commonly used in PTSD research. Posttraumatic stress disorder symptoms specified to the recent childbirth event were assessed at ∼10 days (mean=12.42, standard deviation=2.79), a time point prior to the minimum duration required for a PTSD diagnosis, and again at ∼2 months postpartum (mean=66.24, standard deviation=5.23) using the PTSD Checklist for DSM-5 (PCL-5)^7^, validated against clinician diagnostic assessment^7^. Postpartum depression symptoms were measured (at ∼10 days) with the Edinburgh Postnatal Depression Scale (EPDS). Obstetric complications, including unscheduled or emergency Cesarean delivery and other indicators of maternal and fetal acuity, such as fetal intolerance, postpartum hemorrhage, and placental abruption, were extracted from electronic medical records. The study was approved by the Mass General Brigham IRB.

## Results

Multi-path mediation model examined pathways linking obstetric complications to childbirth-related PTSD symptoms across the early postpartum period (Figure 1). Structural equation modeling (SEM) was used with *lavaan* package in R. Significance was estimated using bias-corrected bootstrap analysis (5,000 resamples). Missing data (9.5% total) were consistent with missing completely at random (Little’s test, p = 0.60) and handled using full information maximum likelihood estimation procedure for missing data on mediators and outcome variable. Analyses were conducted in R (version 4.3.2).

**Figure 1.**
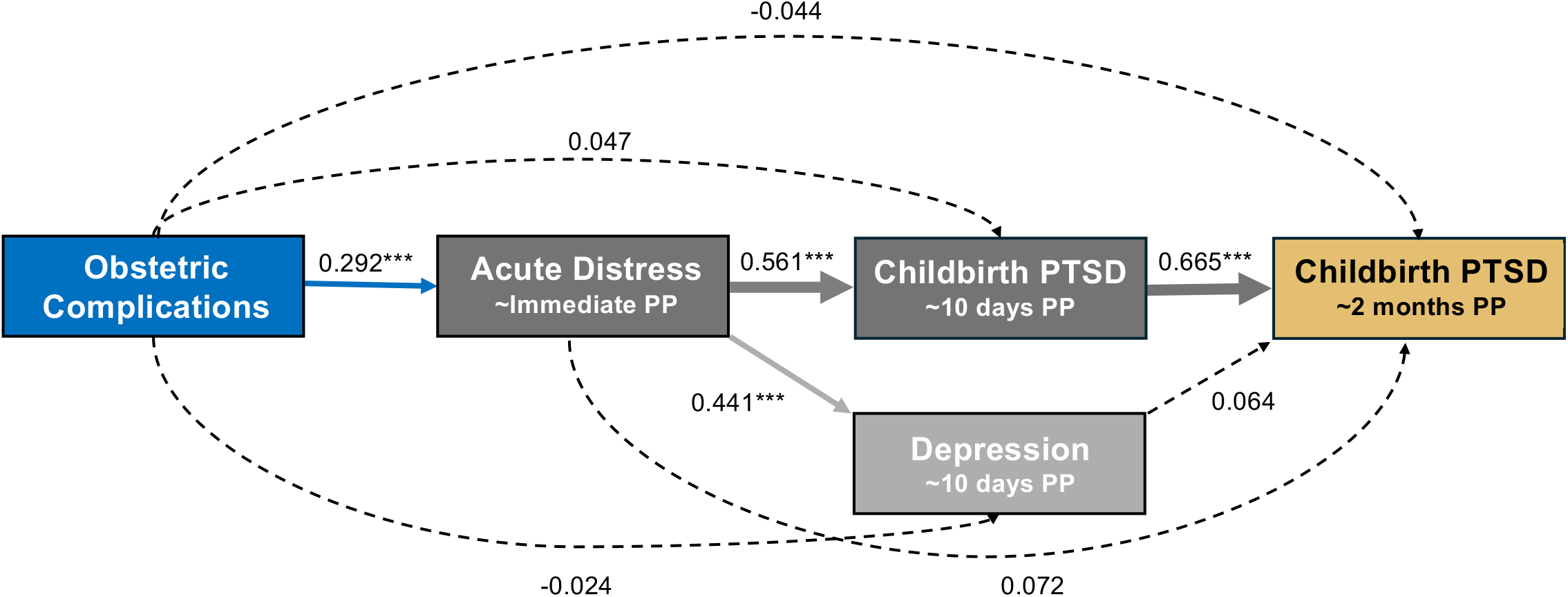
Serial-parallel-multiple-mediator model depicting pathways from obstetric complications to acute distress during childbirth and subsequent trauma-related symptoms across the early postpartum period. Structural equation modeling was used to estimate mediation pathways. Solid arrows indicate statistically significant paths, and dashed arrows indicate non-significant direct paths. Values shown are standardized path coefficients. Indirect effects were tested using bias-corrected bootstrapped confidence intervals (total indirect effect across all mediators, β = 0.168, p < 0.001). Obstetric complications based on electronic medical records. Acute distress was primarily assessed during the postpartum hospitalization using the Peritraumatic Distress Inventory (PDI), and trauma-related symptoms were assessed at approximately 10 days postpartum and again at approximately 2 months postpartum using the PTSD Checklist for DSM-5 (PCL-5). PP = postpartum period. p-value: *** < 0.001.

The mediation model (near-saturated, df=3) revealed that obstetric complications were associated with higher acute distress during childbirth (β = 0.292, p < 0.001) (Fig 1). Acute distress strongly predicted childbirth-related PTSD symptoms at ∼10 days postpartum (β=0.561, p<0.001), which in turn predicted symptom severity at ∼2 months postpartum (β=0.665, p<0.001). The direct association between obstetric complications and PTSD symptoms at 2 months postpartum was not significant, accounting for mediators (β=-0.044). Indirect effects revealed a significant serial mediation pathway through acute distress and early PTSD symptoms (β=0.109, p<0.001). Depression did not mediate these associations.

## Discussion

The immediate postpartum period following delivery provides optimal access to nearly all postpartum patients. Identifying early indicators of risk for childbirth-related PTSD may enable targeted follow-up among women at higher risk and help distinguish PTSD from other maternal psychiatric morbidities, such as postpartum depression.

Our findings suggest that obstetric complications increase risk for later PTSD symptoms indirectly through heightened acute subjective distress during delivery and early postpartum trauma-related symptoms. These findings highlight the importance of incorporating subjective experiences of childbirth into risk assessment, alongside traditional obstetric indicators^4,5^.

These results support a two-step screening approach that can be integrated into existing obstetric care workflows: first, identifying women with elevated acute distress during the immediate postpartum hospitalization, and second, reassessing trauma-related symptoms in the early ^5^postpartum period before symptoms meet the duration criteria for a PTSD diagnosis. Because postpartum mental health screening typically prioritizes depression ^8^, incorporating brief screening for traumatic stress alongside existing approaches may improve identification of women at elevated risk for childbirth-related PTSD and facilitate earlier supportive and preventive care.

## Data Availability

All data produced in the present study are available upon reasonable request to the authors

